# Cardiomyopathy-Associated Mutations in a Hotspot Region at the C-terminal Part of Desmin Coil-2 Domain Impair the Intermediate Filament Assembly

**DOI:** 10.64898/2025.12.15.25342325

**Authors:** Jonas Reckmann, Hendrik Milting, Sabrina Voß, Marco T. Radukic, Franziska Klag, Franziska Flottmann, Alexander Lütkemeyer, Joline Groß, Anna Gärtner, Sandra Landwehr, Dario Anselmetti, Annika Hoyer, Kristian M. Müller, Jan Gummert, Volker Walhorn, Andreas Brodehl

## Abstract

**Background:** The *DES* gene encodes the intermediate filament protein desmin, which connects different multi-protein complexes like the cardiac desmosomes and is highly important for the structural integrity of cardiomyocytes. Pathogenic *DES*-mutations cause filament assembly defects leading to cardiomyopathies. However, most *DES*-variants listed in genetic disease databases are currently classified as variants of unknown significance. Here, we functionally characterized 21 different *DES*-variants of unknown significance and 18 additional proline variants, localized in a highly conserved stretch at the C-terminus of the desmin coil-2 subdomain.

**Methods:** We inserted desmin variants via site-directed-mutagenesis and investigated the filament assembly in transfected cell lines and cardiomyocytes derived from induced pluripotent stem cells by confocal microscopy. In addition, we purified recombinant wild-type and mutant desmin and analyzed the filament formation by atomic force microscopy. Co-expression with wild-type desmin delivered by adeno-associated virus was used to model the heterozygous status of cardiomyopathy patients.

**Results:** Twelve *DES*-variants of unknown significance formed cytoplasmic aggregates, which was likewise verified by atomic force microscopy. Of note, these twelve variants disturb the filament assembly even when co-expressed with wild-type desmin. Using a proline screen, we showed that proline residues localized at nearly each of the positions in this stretch cause filament assembly defects. By modelling the tetrameric structure of desmin, we demonstrated that specific heptad positions as well as positions of intra- and intermolecular ion bridge sites are particularly susceptible mutations that promote desmin aggregation.

**Conclusion:** In summary, our study demonstrated that the highly conserved stretch at the C-terminus of the coil-2 subdomain is a hotspot region, where several pathogenic *DES*-mutations cause an aberrant desmin aggregation. Based on our functional data we suggest to re-classify the aggregate-forming variants as likely-pathogenic mutations rather than variants of unknown significance. Our study may have relevance for the genetic counselling of cardiomyopathy patients with similar *DES*-variants.

## 1. Introduction

Pathogenic *DES* (OMIM, *125660) mutations cause different cardiac and skeletal myopathies including dilated and arrhythmogenic cardiomyopathy ^1–4^. Most pathogenic *DES* mutations are missense variants or small in-frame deletions ^5^. The *DES* gene encodes the muscle-specific type 3 intermediate filament protein desmin ^6^. Desmin filaments connect several different cellular structures like the cardiac desmosomes ^7^, costameres ^8^, Z-band ^9^ and mitochondria ^10^. Therefore, desmin is highly important for the structural integrity of cardiomyocytes ^11^. As a consequence, pathogenic mutations in the *DES* gene compromise the sarcomere structure, which is critical for the contractile function of cardiomyocytes ^12^.

Desmin consists of non-helical head and tail domains and a highly conserved rod domain (Figure 1). The rod domain is composed of two alpha-helical subregions (coil-1 and −2) and mediates the coiled-coil formation ^13,14^. Two desmin dimers form anti-parallel tetramers, which anneal laterally into unit-length filaments (ULF). ULFs are the building blocks of intermediate filaments (IFs) and have a length of ∼60 nm ^15^. The exact molecular structure of the desmin ULFs and desmin filaments are currently unknown. However, Eibauer *et al.* described recently the complete molecular architecture of vimentin filaments using cryo-electron microscopy ^16^. In this model, the alpha-helices form five protofilaments and the non-helical head domains form an amyloid-like structure in the center of the vimentin ^16^.

**Figure 1.**
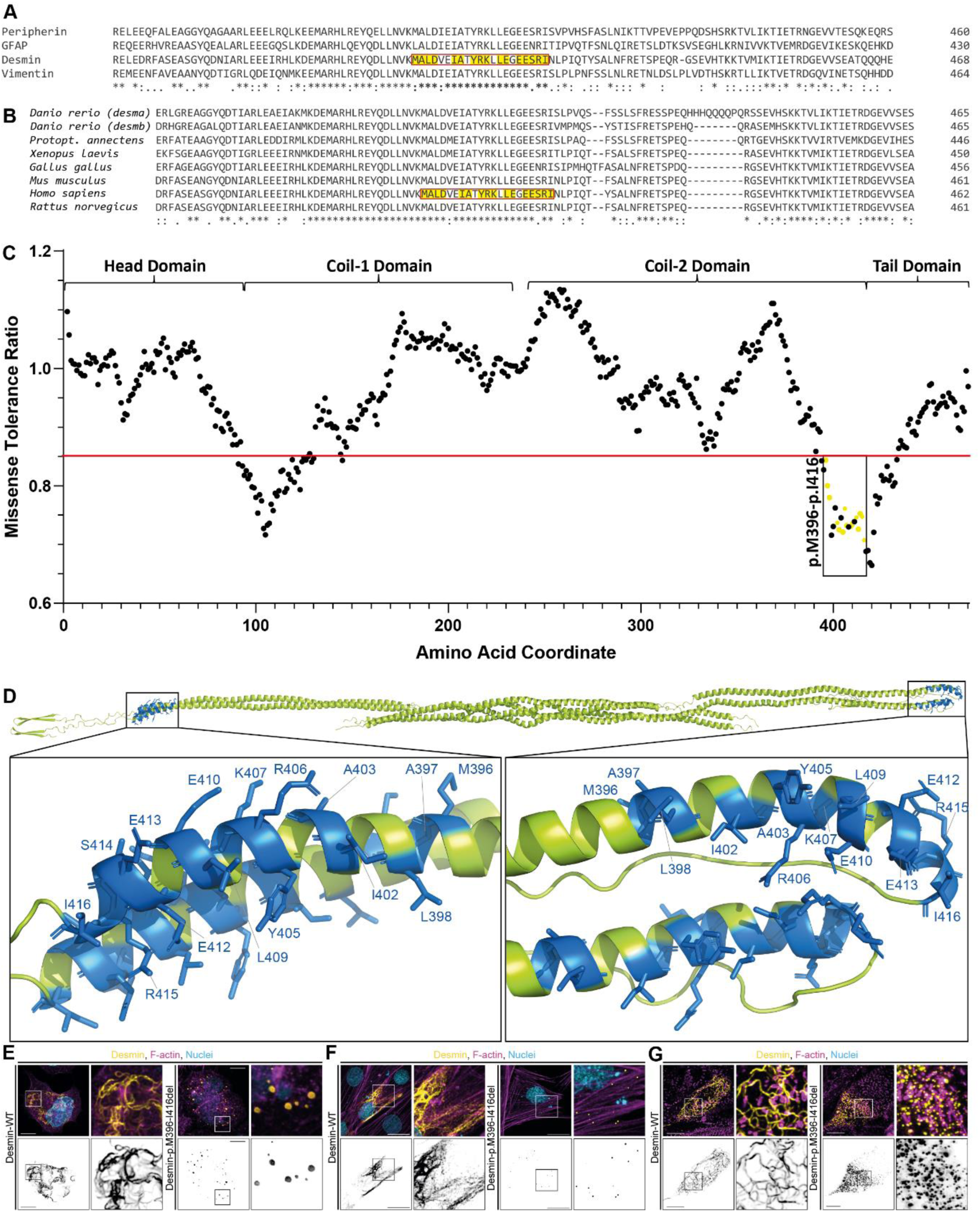
Schematic overview about the highly conserved C-terminal stretch of desmin. **(A)** Partial protein sequence alignment of the human type-3 intermediate filament proteins peripherin (NP_006253.2); glial fibrillary acidic protein, (GFAP, NP_002046.1); desmin (NP_001918.3) and vimentin (NP_003371.2). **(B)** Partial protein sequence alignment of desmin from vertebrate species (NP_001918.3, NP_071976.2, NP_034173.1, NP_571038.2, NP_001070920.2, NP_001080177.1, NP_001383608.1, XP_043931805.1) **(A-B)** A highly conserved stretch at the N-terminus of the coil-2 subdomain is highlighted by a red box. Positions of variants with unknown significance according to the ClinVar database (https://www.ncbi.nlm.nih.gov/clinvar/, October 2025) within this stretch are highlighted in yellow. Identical amino acids within the alignments are labelled with asterisk and similar ones with colons and weak similarities are indicated with dots. **(C)** Missense tolerance ratio (MTR) across the coding region of human desmin. Data were received from the RGC Million Exome Variant Browser (https://rgc-research.regeneron.com/me/gene/DES, October 2025) ^61^. The positions of the different domains are indicated with curly brackets. Lower MTR values (<0.85) indicate regions under selection against missense variants. The highly conserved stretch at the C-terminus of the coil-2 domain is marked by a black box and positions, where variants of unknown significance are known, are shown in yellow. **(D)** Structural overview of the antiparallel desmin tetramer. The positions of variants with unknown significance are shown in marine blue. Representative fluorescence images of SW-13 **(E)** and H9c2 cells **(F)** and iPSC-derived cardiomyocytes **(G)**, which are transfected with wild-type desmin or an artificial desmin deletion (p.M396-I416del) are shown. Desmin is shown in yellow in the upper row and in black in the lower row. F-actin and the nuclei are shown in magenta or cyan. Scale bars represent 10 µm.

Recent findings have revealed a mutation hotspot within the N-terminal segment of the desmin 1A subdomain, where pathogenic variants interfere with IF assembly ^17,18^. Of note, mutations in this genetic hotspot cause severe cardiomyopathies ^19–22^. The 1B subdomain, does not contain such a linear hotspot ^23^. Several proline exchanges or different small in-frame deletions spread over the complete 1B domain affect the filament assembly and cause cardiomyopathies and myopathies ^23–26^. By contrast, currently more than 130 variants of unknown significance (VUS) localized in the coil-2 sub-domain are listed in the ClinVar (https://www.ncbi.nlm.nih.gov/clinvar/) or the Human Gene Mutation Database (https://www.hgmd.cf.ac.uk/).

In this study, we focussed on the functional characterization of 21 different *DES* variants located within a highly conserved C-terminal region of desmin’s coil-2 subdomain, prompted by a notably decreased missense tolerance ratio in the human population, which implies potential pathogenicity (Figure 1A-D). Our functional data revealed that this region is an additional critical hotspot for (cardio)myopathy-associated *DES* mutations.

## 2. Material and Methods

All supporting data are included within the article and its supplemental material. The authors make their data, analytic methods and study material available to other researchers. The generated plasmids can be received from the corresponding author upon reasonable request.

### 2.1 Analysis of Genetic Disease Databases

21 VUS or variants with conflicting classification of pathogenicity listed in the ClinVar (https://www.ncbi.nlm.nih.gov/clinvar/) ^27^ and the Human Gene Mutation Database (https://www.hgmd.cf.ac.uk/) ^28^ were included (December 2024).

### 2.2 Site-Directed Mutagenesis and Plasmid Generation

The plasmid generation of pEYFP-N1-DES (Supplemental Material, Figure S1) was previously described ^29^. The Phusion High-Fidelity DNA Polymerase (Thermo Fisher Scientific, Waltham, USA) was used in combination with appropriate oligonucleotides (Supplemental Material, Table S1) to fuse the *DES* cDNA with *Nco*I and *Bam*HI restriction sites. Subsequently, the polymerase chain reaction (PCR) product was purified by agarose gel electrophoresis combined with the GeneJET PCR Purification Kit (Thermo Fisher Scientific) and was cloned into pET28 (#178901, Addgene, Watertown, USA). Since the *Nco*I restriction site at the 5’-end introduce the missense variant p.S2G, this exchange was re-corrected by site-directed mutagenesis using the Q5 Site-Directed Mutagenesis Kit (New England Biolabs, Ipswich, USA) and appropriate oligonucleotides (Supplemental Material, Table S1). In addition, all other missense or small in-frame deletion variants were introduced into pEYFP-N1-DES and pET28-DES (Supplemental Material, Figure S2) plasmids using the Q5 Site-Directed Mutagenesis Kit (New England Biolabs, Ipswich, USA) or the QuikChange Lightning Kit (Agilent Technologies, Santa Clara, USA) using appropriate oligonucleotides (Supplemental Material, Table S1). The AAV transfer plasmid pZMB1104 (Supplemental Material, Figure S3) encodes a Desmin-mRuby fusion protein controlled by a CMV promoter and flanked by full-length inverted terminal repeats (ITRs) for enhanced adeno-associated virus (AAV) packaging. It was generated by PCR amplification of a *DES*-mRuby gene from pmRuby_N1_DES_WT and restriction cloning with *Mlu*I and *Spe*I (New England Biolabs) into the full-ITR transfer plasmid pZMB0990. Plasmid pZMB1104 was propagated in *E. coli* JW0387-KC at 42 °C as previously described ^30^. Sanger sequencing was used to verify the desmin encoding parts of all plasmids (Macrogen, Amsterdam, Netherlands; Supplemental Material, Table S1 and Figures S4-S6).

### 2.3 Cell Culture

SW-13 and H9c2 cells (American Type Culture Collection, ATCC, Manassas, USA) were cultured in Dulbecco’s Modified Eagle Medium (Invitrogen, Waltham, USA) supplemented with 10 % fetal bovine serum (FBS) and penicillin/streptomycin at 37 °C, 5 % CO_2_. Human induced pluripotent stem cells (iPSCs) were cultured on vitronectin-coated cell culture plates in Essential 8 medium (Thermo Fisher Scientific) as previously described ^21^. Differentiation of iPSCs into cardiomyocytes, selection and maturation was done as previously described ^31^.

### 2.4 Production and purification of adeno-associated virus

AAVs were produced by triple plasmid transfection of HEK-293 cells using the calcium phosphate transfection method. Plasmid-DNA for transfection was prepared using the NucleoSnap Plasmid Midi kit (Macherey-Nagel, Düren, Germany). Plasmids for transfection were pZMB0088 encoding adenoviral helper functions, plasmid pZMB0916 encoding the AAV2 Rep and Cap genes, alternatively plasmid pZMB504 encoding AAV2 Rep and AAV9 Cap, and pZMB1104 encoding the DES-mRuby transgene flanked by ITR for viral packaging. One day before transfection, HEK-293 cells were seeded in 5×15 cm cell culture dishes (Sarstedt, Nümbrecht, Germany), 7.5×10^6^ cells per dish in 15 ml DMEM (Thermo Fisher Scientific), 10% fetal bovine serum (Merck, Darmstadt, Germany), 1 % penicillin/streptomycin (Thermo Fisher Scientific), 37 °C, 5% CO_2_. The next day, 37.5 μg of plasmid-DNA per dish in a molar 1:1:1 ratio of plasmids (pZMB0088:pZMB0916:pZMB1104) was added to 1.25 mL 0.3 M CaCl_2_ and vortexed. The solution was then added dropwise to 1.25 mL 2xHBS (50 mM HEPES, 1.5 mM NaH_2_PO_4_, 280 mM NaCl, pH 7.05) and vortexed again. This transfection mix was added to the cell culture dishes and the cells were incubated for three additional days.

Cells were harvested with a silicon cell scraper (Sarstedt), collected in their respective media, and centrifuged (3000×g, 5 min). The supernatant was discarded, and the pellet was resuspended in 1 mL lysis buffer per dish (50 mM Tris, 150 mM NaCl, 2 mM MgCl_2_, pH 8.0). Three freeze-thaw cycles (−80 °C to 37 °C) were used to lyse the cells. The supernatant was cleared (21,000×g, 5 min), filtered (0.45 µm) and used for batch affinity purification with Poros CaptureSelect AAVX (Thermo Fisher Scientific). 300 µL of affinity resin was incubated with the cleared lysate on a tube roller (10 min). Afterwards, the resin was pelleted (5,000 × g, 5 min) and the supernatant was exchanged with PBS, 0.05% Tween 20 (Carl Roth). After further incubation and pelleting, AAVs were eluted with five resin volumes of 100 mM citric acid, pH 2.0, 5 min, and neutralization with 1 M Tris, pH 8.8. The neutralized eluate was aliquoted and stored at −80 °C until use.

Purified viral vectors were quantified by quantitative PCR. The vector stock was diluted with nuclease-free water containing 0.05% Pluronic F-68 and measured against a standard curve of the vector plasmid pZMB1104 with GoTaq qPCR master mix (Promega, Madison, USA) on a LightCycler480II instrument (Roche, Rotkreuz, Switzerland). Primers were designed to target the CMV promoter on the vector plasmid and were 5’-GGGACTTTCCTACTTGGCA-3’ and 5’-GGCGGAGTTGTTACGACA-3’, obtained in HPLC-grade from Merck. qPCRs had an efficiency of >1.85.

### 2.4 Cell Transfection and Transduction

SW-13 and H9c2 cells were cultured in µ-Slide chambers with glass bottom (ibidi, Gräfelfing, Germany) at the day of transfection. Lipofectamin 3000 (Thermo Fisher Scientific) was used for cell transfections with desmin expression plasmids (ration 1:3) according to the instructions of the manufacturer. The transfected cells were culture for 1 day under standard conditions (37 °C and 5 % CO_2_). For co-transfection experiments SW-13 cells were transduced with AAV9-DES-WT-mRuby using a multiplicity of infections (MOI) of 100.000 and were directly co-transfected with 200 ng of the pEYFP-N1-DES expression plasmids using Lipofectamin 3000 (ratio 1:3). iPSC-derived cardiomyocytes were transfected with 750 ng plasmid per well.

### 2.5 Fixation and Fluorescence Staining of Transfected Cells

One day after transfection, the cells were washed several times with Dulbecco’s Phosphate Buffered Saline (DPBS, Life Technologies, Warrington, UK). Cell fixation was done using 4 % Histofix (Carl Roth, Karlsruhe, Germany) for 15 min at room temperature (RT). Afterwards, the cells were washed with DPBS and were permeabilized using 0.1 % Triton X-100 for 15 min at RT. F-actin was stained using phalloidin conjugated with Texas Red (1:400, Thermo Fisher Scientific) for 40 min at RT. After washing with DPBS, 4′,6-diamidino-2-phenylindole (DAPI, 1 µg/mL) was used for 5 min at RT for staining of the nuclei. Finally, the cells were washed several times with DPBS.

### 2.6 Confocal Microscopy Analysis

As previously described in detail, the TCS SP8 system (Leica Microsystems, Wetzlar, Germany) was used for confocal laser scanning microscopy ^17^. 3D stacks of the transfected cells were generated and were presented as maximum intensity projections using the Las X software (Leica Microsystems). The Huygens Essential software (Scientific Volume Imaging, Hilversum, Netherlands) was used for deconvolution and the Fiji software (Version 1.54p) ^32^ was used in combination with EzColocalization plugin ^33^ for colocalization analysis.

### 2.7 Expression and Purification of Recombinant Desmin

*Escherichia coli* (One Shot BL21 DE3, Thermo Fisher Scientific) were transformed with pET28-DES expression plasmids by heat shock (30 sec, 42 °C). The transformed bacteria were selected using kanamycin (50 µg/mL) over night on agar plates (37 °C). After picking single colonies, the bacteria were cultured at 37 °C in LB medium (Carl Roth) under intensive shaking. When the OD_600_ reached a value of >0.6, isopropyl β-D-1-thiogalactopyranoside (IPTG, 1 mM) was added to the bacteria suspension. 4 h after induction of desmin expression, the bacteria were collected by centrifugation (30 min, 2.500 × g, 4 °C). The bacteria pellets were stored at −80 °C until purification of recombinant desmin.

Frozen pellets were treated with DNase and lysozyme for isolation of the inclusion bodies (IBs). Afterwards, the isolated IBs were transferred under denaturing conditions to ion-exchange chromatography. HiTrap DEAE FF columns (Cytiva, Marlborough, USA) were equilibrated with binding buffer (8 M urea, 10 mM Tris-HCl, pH 8.0). Desmin molecules were eluted by a linear salt gradient (8 M urea, 10 mM Tris-HCl, 1 M NaCl, pH 8.0). Afterwards, the samples were pooled and loaded to a HisTrap FF columns (Cytiva). The columns were equilibrated (8 M urea, 20 mM NaH_2_PO_4_, 20 mM imidazole, pH 7.4) and purified desmin eluted using elution buffer (8 M urea, 20 mM NaH_2_PO_4_, 500 mM imidazole, pH 7.4). Afterwards, buffer was exchanged against storage buffer (8 M Urea, 20 mM Tris-HCl, 300 mM imidazole, pH 7.4) and the recombinant desmin was stored at −80 °C until use for AFM analysis.

### 2.8 Desmin Preparation and Atomic Force Microscopy Analysis

Recombinant desmin filament assembly for atomic force microscopy (AFM) was followed as previously described ^29^. In brief, a stepwise dialysis (1 h, room temperature) in DP-buffer (5 mM Tris-HCl, 1 mM dithiothreitol, pH 8.4) was performed to reduce the urea. Recombinant desmin filament formation was induced by addition of sodium chloride (200 mM NaCl, 45 mM Tris-HCl, pH 7.0) for 1 h at 37 °C. Ten microliter of desmin samples were immobilized on freshly cleaved MICA substrates (Plano, Wetzlar, Germany). After short incubation time (∼ 1 min) the samples were rinsed off with deionized water and gently dried under nitrogen flow. Topographical images in ambient conditions were taken in tapping mode with Tap300AI-G cantilever (Budget Sensors, Sofia, Bulgaria) with a JPK Nanowizard system (JPK Bruker, Berlin, Germany). Otherwise, preassembled desmin was applied in a chamber for measurements in buffer (200 mM NaCl, 45 mM Tris-HCl, pH 7.0) using USC-F0.3-k0.3 cantilever (Nano World, Neuchâtel, Switzerland). Imaging analysis was done with Gwyddion software (version 2.69, https://gwyddion.net/).

### 2.9 Molecular Modelling and Protein Alignment

As previously described ^34^, we used the molecular structure of the homologous protein vimentin ^16^ for homology modeling using the SWISS-MODEL server ^35^. PyMOL Molecular Graphics System (Version 3.1.6.1, Schrödinger, New York, USA) was used for visualization of the desmin structure. The Clustal Omega Multiple Sequence Alignment tool (https://www.ebi.ac.uk/jdispatcher/msa/clustalo) ^36^ was used for protein alignments.

### 2.10 Statistical Analysis

Statistical analysis was performed to evaluate the risk for the mutations of forming aggregates. As discrete outcome variable we used the number of cells which revealed the presence of aggregates for each mutation, ranging between 0 and 100 cells. For each mutation the experiment was repeated four times, resulting in four independent observations concerning the number of cells. The mutation variants with the wild-type as reference level were used as independent variable. Due to underdispersion, detected via a dispersion test, a generalised Poisson regression model was applied. As a result, we obtain for each mutation the relative risk (with 95%-CI) for showing aggregates compared to the wild-type. All statistical analyses were performed using R version 4.5.2.

## 3. Results

Different type-3 IF proteins like vimentin, peripherin, glial fibrillary acidic protein (GFAP) and desmin show high homology of their rod domains. Especially, the N-terminal part of the coil-1 and the C-terminal part of the coil-2 subdomains are highly homologous between different type-3 IF proteins (Fig. 1A) and are likewise highly conserved between different vertebrate species (Fig. 1B). Of note, the missense tolerance ratio (MTR) is reduced in these two regions (Fig. 1C). Recently, we have shown that the N-terminal part of coil-1 subdomain is a critical hotspot region, where several pathogenic cardiomyopathy-associated mutations affect the filament assembly ^18,37^. However, 21 different VUS or variants with conflicting classification at the C-terminus of the coil-2 subdomain are currently listed in the ClinVar database (https://www.ncbi.nlm.nih.gov/clinvar, October 2025, Fig. 1D). Since it is known, that pathogenic cardiomyopathy-associated *DES* mutations in this region disturb the desmin filament assembly ^38,39^, we proposed that functional characterization can help to reclassify these VUS.

According to the ACMG guidelines ^40^, we have validated the aberrant aggregate formation using eleven known pathogenic *DES* variants (p.E108K, p.K109N, p.L115I, p.N116D, p.Y122H, p.Y122S, p.R127P, p.A337P, p.R350P, p.E401D, p.R406W) and one negative variant (p.A213V ^41^, Fig. S7). To analyse the relevance of the highly conserved C-terminal stretch, we generated a deletion ranging from p.M396 to p.I416 (p.M396-p.I416del). Wild-type desmin formed in both cell lines and in cardiomyocytes derived from induced pluripotent stem cells intermediate filaments (Fig. 1E-G), while the artificial deletion mutant desmin-p.M396-p.I416del formed aberrant cytoplasmic desmin aggregates (Fig. 1E-G and Supplemental Material, Figure S8). Relative risks with 95%-CI for each mutation forming aggregates in SW-13, H9c2 cells and cardiomyocytes – compared to wild-type – are given in Supplemental Material, Tables S3-S5. SW-13 cells were used since they do not express desmin or any other endogenous cytoplasmic IF protein ^42^. H9c2 cells are cardiac myoblasts and express endogenous desmin ^43^. These findings underline the general impact of the highly conserved stretch localized at the coil-2 subdomain of desmin for the filament formation.

To evaluate the impact of these VUS, we generated 21 different expression plasmids encoding these VUS and analyzed the filament or aggregate formation in SW-13, H9c2 cells and in cardiomyocytes derived from induced pluripotent stem cells. Nine VUS (p.M396T, p.A397S, p.A397T, p.I402T, p.A403D, p.K407Q, p.S414G, p.R415W and p.I416M) formed desmin filaments, comparable to wild-type desmin although they have high AlphaMissense or REVEL scores (Fig. 2). In contrast, twelve VUS (p.L398P, p.I402S, p.Y405H, p.R406L, p.R406Q, p.L409P, p.E410K, p.E412K, p.E413G, p.R415G and p.R415Q) caused aberrant cytoplasmic desmin aggregates (Fig. 2) similar to the desmin deletion p.M396-p.I416del (Fig. 1E-F) and pathogenic controls (Supplemental Material, Figure S7).

**Figure 2.**
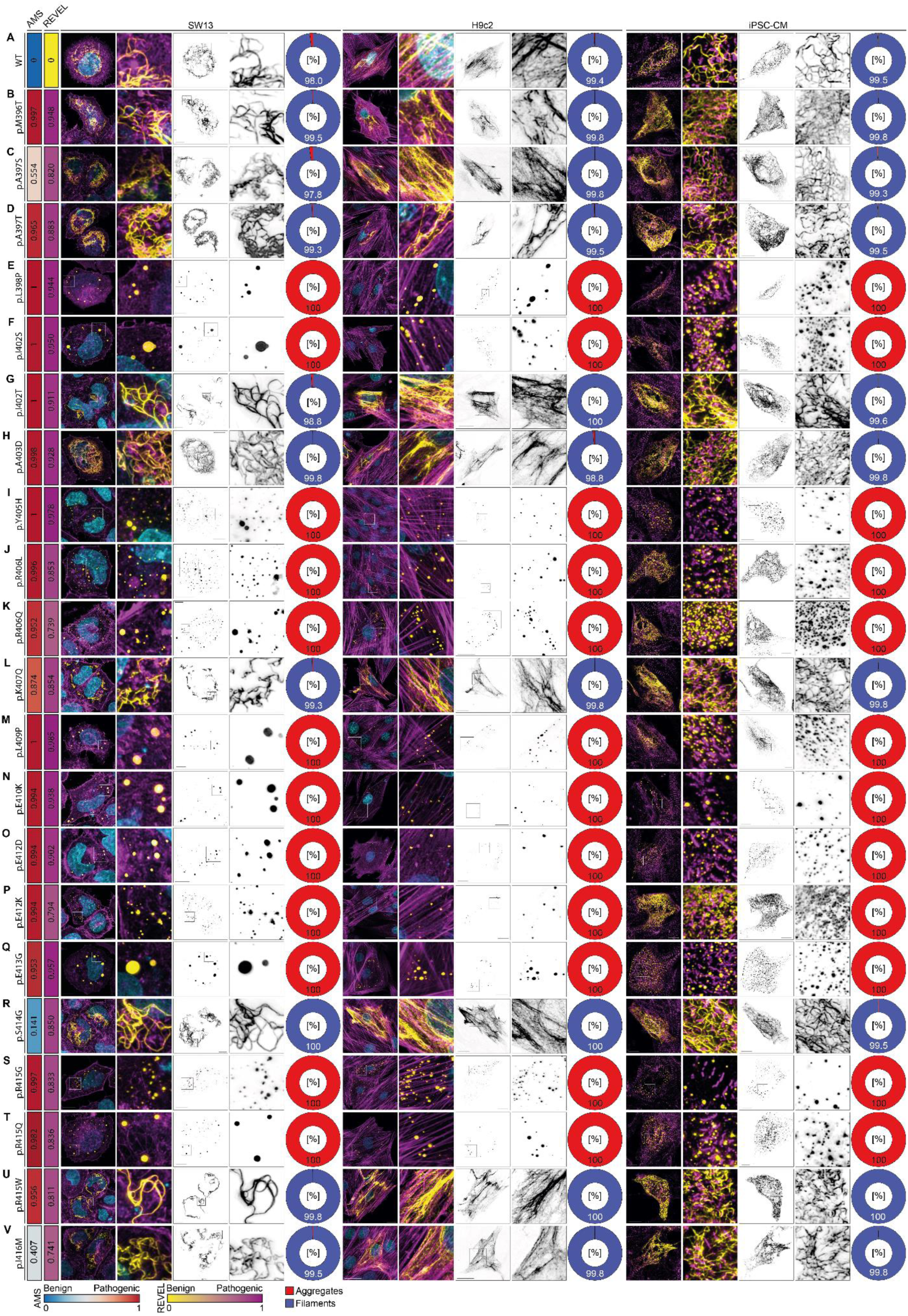
Functional analysis of wild-type desmin and missense variants in cell culture. SW-13, H9c2 cells and iPSC-derived cardiomyocytes were transfected with either wild-type or mutant desmin plasmids. Representative maximum intensity projections are shown. Desmin-EYFP appears in yellow or black, F-actin and α-actinin-2 are shown in magenta and nuclei in cyan. Scale bar corresponds to 10 µm as well as the highlighted square. For each missense variant, two prediction scores from AlphaMissense Pathogenicity (AMS) and Rare Exome Variant Ensemble Learner (REVEL) are presented as heat maps on a 0-1 scale. The percentages of different cellular phenotypes (filaments, blue; aggregates, red) are illustrated by pie charts.

To verify the cell culture data in a second approach, we purified recombinant wild-type and mutant desmin and analyzed the structure of the assembled desmin species by AFM. Wild-type desmin formed filaments of different lengths (Fig. 3A and 3a). In contrast, the deletion mutation (p.M396-I416del) and a pathogenic positive control (p.R406W) formed exclusively aberrant small desmin structures (Fig. 3B-C and 3b-c). Comparable to this finding, all other aggregate forming variants (p.L398P, p.I402S, p.Y405H, p.R406L, p.R406Q, p.L409P, p.E410K, p.E412D, p.E412K, p.E413G, p.R415G and p.R415Q) formed small aberrant structures verifying their intrinsic filament assembly defects (Fig.3D-O and 3d-o).

**Figure 3.**
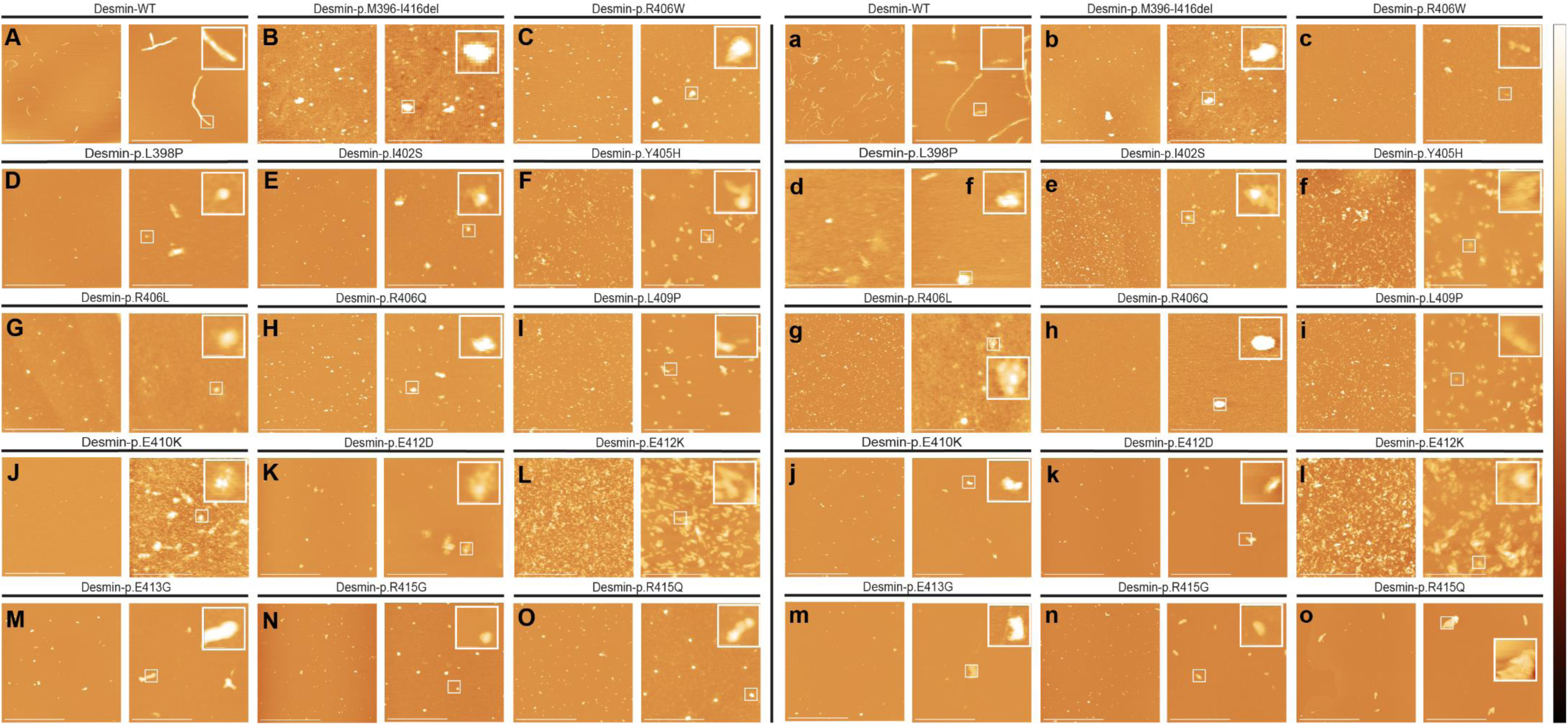
Atomic force microscopy analysis of recombinant wild-type and mutant desmin. Topographical images of wild-type desmin **(A, a)** and 14 aggregate forming variants including the deletion (p.M396-I416del, **B, b**) and the positive pathogenic mutation (p.R406W, **C, c**) analyzed by atomic force microscopy in ambient conditions (**A-O**) and in buffer **(a-o)**. The lateral scale bar corresponds to 5 µm (left image) and 1 µm (right image) including the 100 nm zoom. The colour scale represents a height of 10 nm wild-type and 25 nm for the mutant desmin structures.

Since most patients carry heterozygous *DES* variants, we performed double transfection-transduction experiments of wild-type and mutant desmin conjugated with green (EYFP) and red (mRuby) fluorescence proteins. In the control experiments, wild-type desmin fused to EYFP or mRuby formed filaments consisting of both forms (Fig. 4A). Similarly, the other filament forming variants (p.M396T, p.A397S, p.A397T, p.I402T, p.A4093D, p.K407Q, p.S414G, p.R415W and p.I416M) assembled into mixed filaments consisting of both forms. Of note, the aggregate forming variants (p. L398P, p.I402S, p.Y405H, p.R406L, p.R406Q, p.L409P, p.E410K, p.E412D, p.E412K, p.E413G, p.R415G and p.R415Q) as well as the pathogenic control (p.R406W) and the artificial deletion variant (p.M396T-p.I416del) formed aggregates consisting of wild-type and mutant desmin (Fig. 4), which is in good agreement with a dominant negative inheritance.

**Figure 4.**
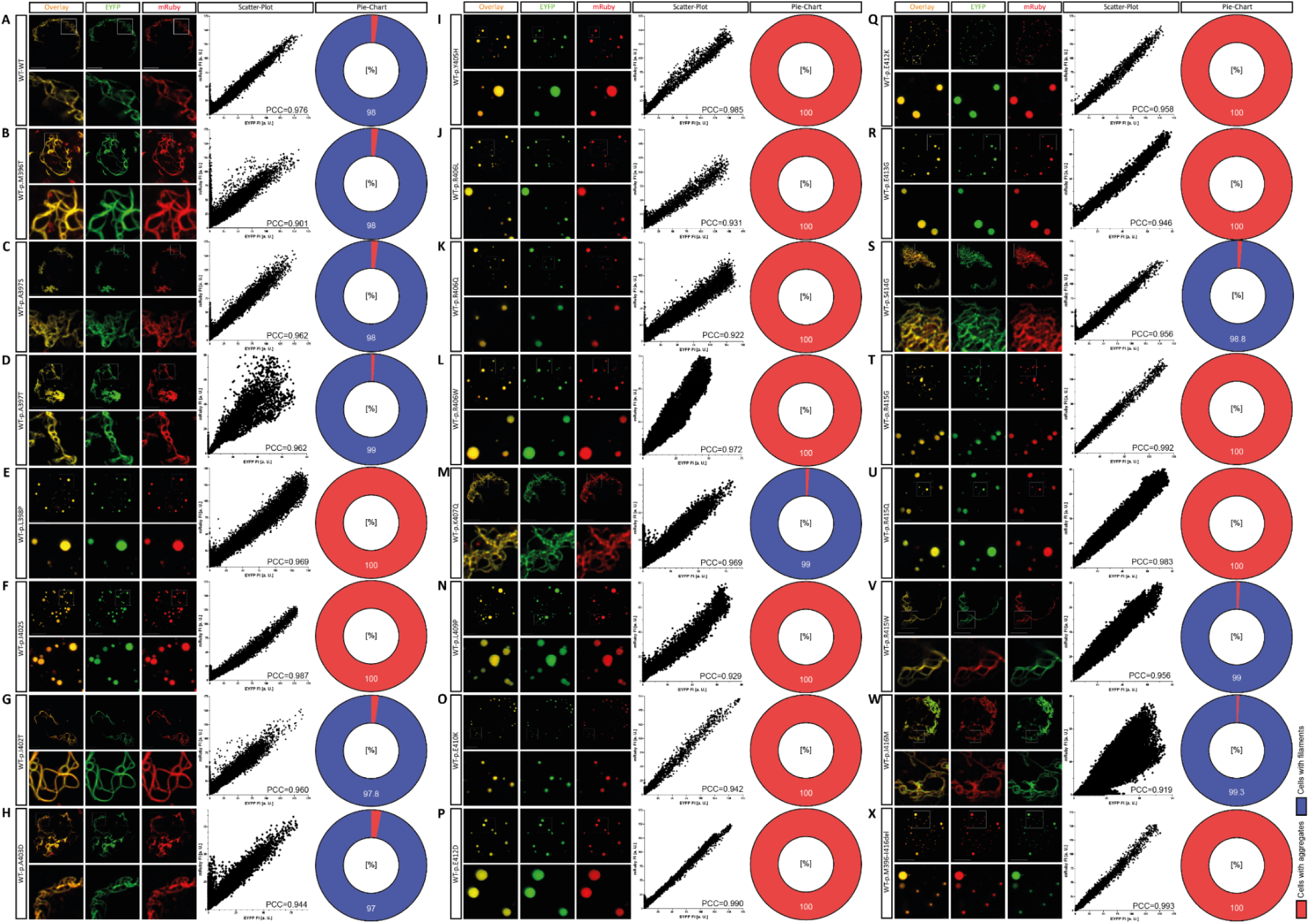
Double transfection experiments. Representative images of SW-13 cells transduced with adeno-associated virus 9 encoding desmin-WT-mRuby (red) and co-transfected with expression pEYFP-N1-DES plasmids encoding wild-type **(A)** or mutant **(B-X)** desmin (green) are shown. Colocalization overlay is shown in yellow. Scale bars represent 10 µm. Representative scatter plots are visualized and were used to calculate the indicated Pearson correlation coefficients (PCC). The percentage of cells with filaments (blue) or aggregates (red) is summarized as pie charts (n=4, >100 transfected cells per independent transfection experiment).

Proline residues may interfere with the desmin filament assembly, since proline is known as a strong breaker of the α-helices. Recently, we showed that proline residues at the a- and d-position within the desmin heptad sequence of the 1B sub-domain disturb the filament assembly ^23^. Systematic data at the conserved C-terminal stretch are currently missing. Therefore, we performed a systematic proline screen, where we introduced proline residues at each position within the stretch (Fig. 5). Four variants (p.M396P, p.A397P, p.V400P and p.I416P) localized at the beginning or end of the stretch formed a borderline phenotype with small filamentous or mixed structure (Fig. 5). All other desmin proline variants (p.L398P, p.D399P, p.E401P, p.I402P, p.A403P, p.T404P, p.Y405P, p.R406P, p.K407P, p.L408P, p.L409P, p.E410P, p.G411P, p.E412P, p.E413P, p.S414P and p.R415P) were unable to assemble into regular filaments (Fig. 5), indicating a high susceptibility of this region for helix destabilizing effects by proline residues.

**Figure 5.**
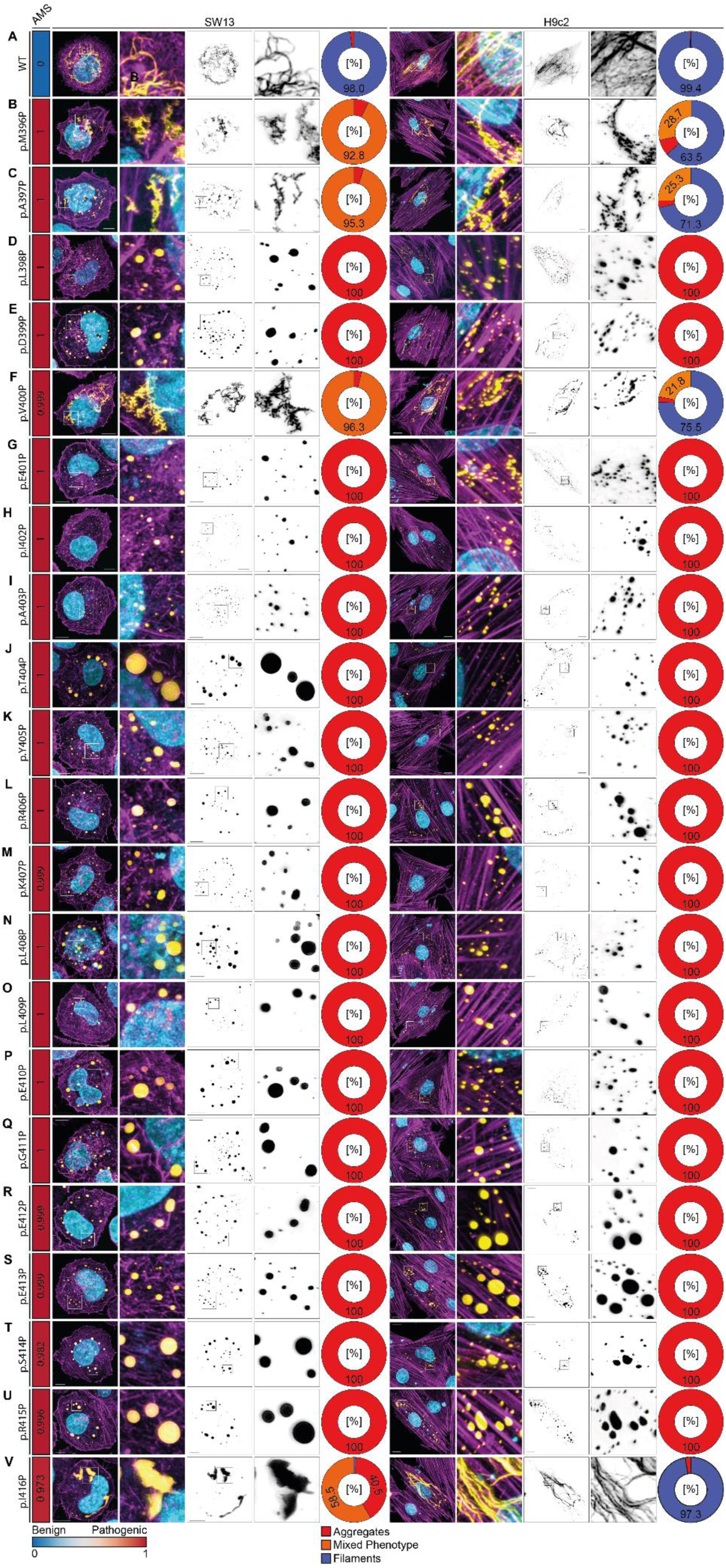
Proline screen of the C-terminal stretch of the coil-2 domain. Representative fluorescence images of SW-13 cells and H9c2 cells expressing wild-type desmin **(A)** or proline variants **(B-V)** are shown. The AlphaMissense prediction scores (AMS) were derived from the AlphaFold Protein Structure Database (https://alphafold.ebi.ac.uk/, October 2025)^62^. Desmin is shown in yellow (or in black, right row), F-actin in magenta and the nuclei in cyan. Scale bars represent 10 µm. The percentage of cells with filaments (blue), aggregates (red) or a mixed phenotype (orange) is shown as pie charts (n=4, >100 transfected cells per independent transfection experiment).

## 4. Discussion

Most pathogenic cardiomyopathy-associated desmin (*DES*) mutations are small in-frame deletions or missense mutations ^5^. However, most of the known *DES* variants listed in genetic disease database like ClinVar or the HGMD are currently classified as VUS, since functional data and co-segregation data are not available. Recently, according to the guidelines of Brnich *et al.* ^40^, we established a robust cell-culture based assay to differentiate between filament- and aberrant aggregate-forming desmin variants ^23^. Using this assay, we functionally characterized over 300 different missense variants localized in the head- ^17^, 1A- ^18^ and 1B domain ^23^ revealing a hotspot region in N-terminal region of desmin, where several mutations affect the desmin filament assembly leading to aberrant cytoplasmic desmin aggregates. In addition, it is known, that several proline residues especially at hydrophobic a- and d-positions within the 1B-domain disturb the desmin filament assembly^23^.

Human population data derived from the Million Exome Variant Browser (https://rgc-research.regeneron.com/me/gene/DES, 25^th^ November 2025) show a decreased missense tolerance ratio of a highly conserved C-terminal region of the coil-2 domain (Fig. 1C). These data indicate that missense variants in this region of desmin are more likely to be deleterious or disease-causing. In good agreement with this hypothesis, cardiomyopathy-associated *DES* mutations (p.E401D and p.R406W) have been previously identified ^39,44^. In addition, Herrmann *et al.* generated at the corresponding position a knock-in mouse model (*Des*-p.R405W), which develops a severe cardiomyopathy in combination with a skeletal myopathy ^38^.

To investigate this interesting desmin region in more detail, we generated first a complete deletion of this region (p.M396-I416del). Desmin-p.M396-I416del caused an aberrant cytoplasmic aggregation as previously described for other pathogenic deletions ^26,45,46^. Of note, 21 different VUS localized in this region are listed in ClinVar (https://www.ncbi.nlm.nih.gov/clinvar/, 5^th^ December 2024). To fill this gap of knowledge, we functionally investigated these 21 different VUS revealing for twelve of them a desmin filament assembly defect (Fig. 6A) in transfected SW-13, H9c2 or hiPSC- derived cardiomyocytes. Nine different desmin variants formed filaments similar to wild-type desmin (Fig. 6A). The AFM analysis supports likewise the filament assembly defect of these mutants. Since most patients carrying *DES* variants present a heterozygous genotype, we performed co-transfection experiments. These experiments revealed for all aggregate-forming mutants a dominant-negative effect on filament assembly (Fig. 4), which is in good agreement with an autosomal dominant-negative inheritance ^47^. On the one hand aggregate forming mutations are localized at positions of the heptad. On the other hand, aggregate forming mutations affect positions, where intra- and intermolecular ion bridges likely occur in the wild-type protein (Fig. 6B-C and Supplemental Material, Figure S9).

**Figure 6.**
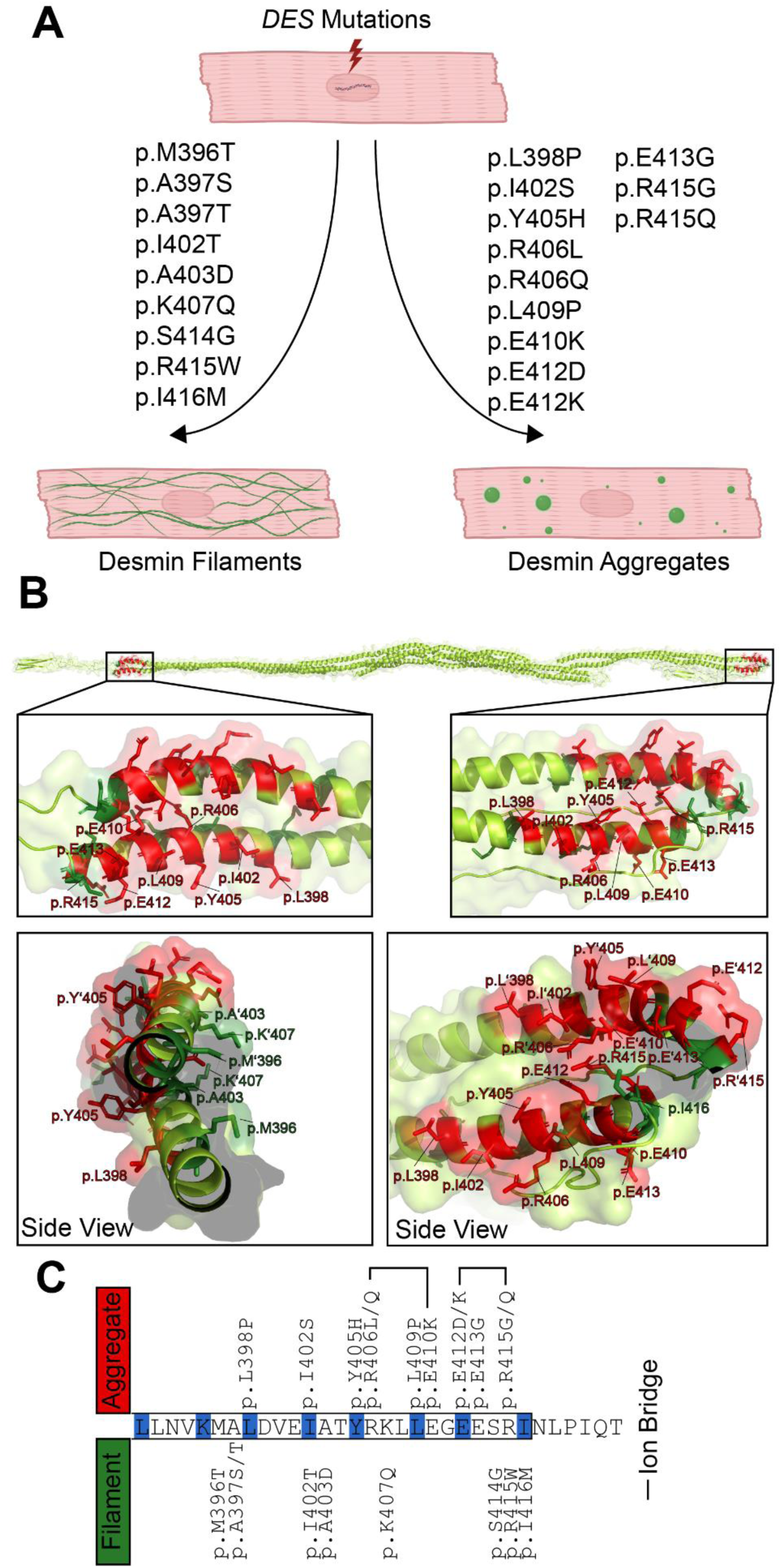
Schematic summary. **(A)** 21 different variants of unknown significance (VUS) in a highly conserved stretch at the C-terminus of the Coil-2 domain of desmin have been investigated. Nine variants formed filaments similar to wild-type desmin, whereas twelve variants formed aberrant desmin aggregates. **(B)** Structural overview of the desmin tetramer. The positions of the aggregates forming variants are shown in red, whereas the positions of filament forming mutants are shown in green. **(C)** Schematic overview of the localization of the VUSs localized at the C-terminus of the desmin coil-2 domain. The heptad is highlighted in blue. Inter- and intramolecular ion bridges are indicated with lines.

Therefore, we suggest to re-classify these aggregate forming variants as likely-pathogenic *DES* variants, which might be relevant for disease classification of similar *DES* variants in the future. Two aggregate-forming *DES* mutations (p.L398P and p.L409P) cause an exchange against proline residues. Proline is known as breaker of α-helices ^48^. Therefore, we addressed in this study, if proline residues localized in the highly conserved C-terminal stretch might be deleterious in general Supplemental Material, Figure S10). These experiments revealed that the insertion of proline residues at nearly all positions within this stretch cause an aberrant cytoplasmic desmin aggregation indicating that proline residues at the C-terminus of the coil-2 subdomain have a severe detrimental effect on filament assembly and may be considered as genetic risk factors for cardiomyopathy-associated *DES* variants.

It is known, that mutations in other paralogous IF-encoding genes like *GFAP* (OMIM, *137780) or *LMNA* (OMIM, *150330) cause Alexander disease (OMIM, #203450) or different (cardio)myopathies (OMIM, #115200) ^49–51^. Interestingly, Alexander-disease causing *GFAP* mutations have been described at the corresponding positions to the investigated desmin variants (*GFAP*-p.E373K ^52^ ↔ *DES*-p.E412K; *GFAP*-p.Y366H ^53^ ↔ *DES*-p.Y405H). Similarly, disease-causing *LMNA* mutations have been described, that correspond to the desmin-aggregate forming mutations (*LMNA*-p.L369P ^54^ ↔ *DES*-p.L398P, *LMNA*-p.R377L ^55^ ↔ *DES*-p.R406L). These data indicate that the highly conserved C-terminal part of the coil-2 subdomain is in general important for the function of different IF proteins.

In summary, we present here functional data, which justify the re-classification of several *DES* missense variants as likely-pathogenic mutations (p.L398P, p.I402S, p.Y405H, p.R406L, p.R406Q, p.L409P, p.E410K, p.E412D, p.E412K, p.E413G, p.R415G and p.R415Q) according the ACMG guidelines ^56^. In addition, our study provides evidence, that the C-terminal part of the coil-2 region is a hotspot area, where several desmin mutations cause filament assembly defects leading to (cardio)myopathies. Therefore, this work may have relevance for the clinical and genetic counselling of patients carrying similar *DES* variants.

## Limitations

Desminopathies are complex and can cause a broad range of cardiac and skeletal myopathies ^57^. Therefore, our study may not fully capture all features of *DES*-associated cardiomyopathies, like *e. g.* impact on the mitochondrial integrity ^58–60^, but focus on the structural filament-assembly defects. In addition, we can not exclude changes in the nanomechanical properties of filament forming VUS in our transfection assay.

## Non-standard Abbreviations and Acronyms

AAV: Adeno-Associated Virus
AFM: Atomic Force Microscopy
DAPI: 4′,6-Diamidino-2-phenylindole
DMEM: Dulbecco’s Modified Eagle Medium
EDTA: Ethylenediaminetetraacetic Acid
EYFP: Enhanced Yellow Fluorescent Protein
FCS: Fetal Calf Serum
HTx: Heart Transplantation
IF: Intermediate Filaments
iPSCs: Induced Pluripotent Stem Cells
ITRs: Inverted Terminal Repeats
DPBS: Dulbecco’s Phosphate Buffered Saline
PCR: Polymerase Chain Reaction
MAF: Minor Allele Frequency
SD: Standard Deviation
ULFs: Unit Length Filaments
VUS: Variant of Unknown Significance

## Author Contributions

Conceptualization (VW, AB); Data curation (JR, SV, MTR, AB); Formal analysis (JR, SV, AB); Statistical Analysis (S.L., A.H.); Funding acquisition (MTR, KM, HM, AB); Investigation (JR, SV, FK, FF, AL, MTR, AB); Project administration (VW, AB); Resources (KM, JG, DA); Supervision (VW, AB); Visualization (JR, VW, AB); Roles/Writing - original draft (JR, AB); and Writing - review & editing (all authors).

## Supplemental Materials

Tables S1-S5.

Figure S1-S10.

